# Precise prostate contours: setting the bar and meticulously evaluating AI performance

**DOI:** 10.1101/2024.10.21.24315771

**Authors:** Yuze Song, Anna Dornisch, Robert T Dess, Daniel JA Margolis, Eric P Weinberg, Tristan Barrett, Mariel Cornell, Richard E Fan, Mukesh Harisinghani, Sophia C. Kamran, Jeong Hoon Lee, Cynthia Xinran Li, Michael A Liss, Mirabela Rusu, Jason Santos, Geoffrey A Sonn, Igor Vidic, Sean A Woolen, Anders M Dale, Tyler M Seibert

**Affiliations:** Department of Radiation Medicine, University of California San Diego, La Jolla, CA, USA; Department of Electrical and Computer Engineering, University of California San Diego, La Jolla, CA, USA; Department of Bioengineering, University of California San Diego, La Jolla, CA, USA; Department of Radiology, University of California San Diego, La Jolla, CA, USA; Department of Urology, University of California San Diego, La Jolla, CA, USA; Department of Radiation Oncology, University of Michigan, Ann Arbor, MI, USA; Department of Radiology, Cornell University, Ithaca, NY, USA; Department of Clinical Imaging Sciences, University of Rochester Medical Center, Rochester, NY, USA; Department of Radiology, University of Cambridge, Cambridge, United Kingdom; Radformation, New York, NY, USA; Department of Urology, Stanford School of Medicine, Palo Alto, CA, USA; Department of Radiology, Massachusetts General Hospital, Boston, MA, USA; Department of Radiation Oncology, Massachusetts General Hospital, Boston, MA, USA; Department of Radiology, Stanford School of Medicine, Palo Alto, CA, USA; Institute for Computational and Mathematical Engineering, Stanford University, Palo Alto, CA, USA; Department of Urology, University of Texas Health Sciences Center San Antonio, San Antonio, TX, USA; Department of Biomedical Data Science, Stanford University, Palo Alto, CA, USA; Quibim, New York, NY, USA; Cortechs.ai, San Diego, CA, USA; Department of Radiology and Biomedical Imaging, University of California San Francisco, San Francisco, CA, USA; Department of Neurosciences, University of California San Diego, La Jolla, CA, USA; Halıcıoğlu Data Science Institute, University of California San Diego, La Jolla, CA, USA

## Abstract

**Introduction:** Evaluation of artificial intelligence (AI) algorithms for prostate segmentation is challenging because ground truth is lacking. We aimed to (1) create a reference standard dataset with precise prostate contours by expert consensus and (2) evaluate various AI tools against this standard.

**Materials and methods:** We obtained prostate MRI cases from six institutions from the Quantitative Prostate Imaging Consortium. A panel of four experts (two genitourinary radiologists, two prostate radiation oncologists) meticulously developed consensus prostate segmentations on axial *T_2_*-weighted series. We evaluated the performance of six AI tools (three commercially available, three academic) using Dice scores, distance from reference contour, and volume error.

**Results:** The panel achieved consensus prostate segmentation on each slice of all 68 patient cases included in the reference dataset. We present two patient examples to serve as contouring guides. Depending on the AI tool, median Dice scores (across patients) ranged from 0.80 to 0.94 for whole prostate segmentation.

For a typical (median) patient, AI tools had a mean error over the prostate surface ranging from 1.3 to 2.4 mm. They maximally deviated 3.0 to 9.4 mm outside the prostate and 3.0 to 8.5 mm inside the prostate for a typical patient. Error in prostate volume measurement for a typical patient ranged from 4.3% to 31.4%.

**Discussion:** We established an expert consensus benchmark for prostate segmentation. The best-performing AI tools have typical accuracy greater than that reported for radiation oncologists using CT scans (most common clinical approach for radiotherapy planning). Physician review remains essential to detect occasional major errors.

## Introduction

Artificial Intelligence (AI) tools are being developed and deployed rapidly, especially for medical imaging^1^. AI tools are valuable additions to clinical medicine in an era of increasingly vast data^2^. However, adverse consequences may follow if AI tools are deployed without proper stewardship by the medical community. Expert medical professionals must set the bar used to judge the performance of AI tools their field will use.

One area of increasing interest is the development of AI algorithms for prostate auto-segmentation, an important task for diagnostic radiologists, radiation oncologists, and urologists. Establishing acceptable accuracy and precision metrics requires knowledge about what constitutes a clinically meaningful error. In diagnostic radiology, the most common segmentation applications are measurement of prostate volume for PSA-density^3,4^, whole gland segmentation for fusion biopsy, and target segmentation for fusion biopsy. The PSA-density task does not require as much accuracy as more difficult applications (measuring extra-prostatic extension risk). While many prostate auto-segmentation tools for MRI are available, most were designed and validated to assist with segmentation for MRI-ultrasound fusion biopsy^5^. It is unknown how well any available auto-segmentation tool works against a true gold standard, as there is no consensus guideline for prostate delineation on MRI. This has particular implications in radiotherapy, where small errors can impact patient outcomes (a randomized trial showed differences in prostate contours as small as 2 mm can measurably increase toxicity)^6^. An improved standard is needed for accuracy for prostate delineation for radiotherapy planning and can also serve as the reference for evaluation of prostate auto-segmentation tools.

Excellent reference standard contours are a prerequisite for a meaningful comparison and properly validated tool. Our objectives are two-fold: 1) create a reference standard dataset with highly accurate prostate contours via expert consensus and 2) meticulously evaluate an array of AI tools for prostate segmentation on MRI against the defined consensus reference standard. In accomplishing the first objective, we present a detailed contouring guide for physician education.

## Methods

### Cases

We obtained patient cases from six institutions from the Quantitative Prostate Imaging Consortium (QPIC) as part of a study of prostate cancer detection approved by the institutional review board at each institution^7,8^. All patients underwent prostate multi-parametric MRI (mpMRI) for known or suspected prostate cancer. We included at least 10 cases from each institution. Cases in the database were selected from each institution in roughly sequential order. We excluded cases with prominent artifacts (hip implants, prominent bowel gas, and/or very large body habitus that interfered with image quality). The dataset was enriched to ensure approximately one-third of cases from each site had a prominent median lobe (prostate extension into the bladder) because this is common in clinical practice, and it is important to understand how automated tools perform for common anatomic variations.

### Reference Standard Contour Development

We convened a panel of four experts to develop the reference standard prostate segmentation dataset. Our panel included two genitourinary (GU) radiologists (with 12 and 21 years’ experience, each surpassing the number of MRIs reported to qualify as prostate MRI experts^9^) and two GU radiation oncologists specializing in MRI-guided prostate radiotherapy (10 years’ experience each). One of the GU radiation oncologists, also a prostate MRI researcher, generated initial contours on high-resolution axial *T_2_*-weighted slices. The panel met weekly from March to June 2024 to review the initial contours slice-by-slice. All three planes were always visualized, and high-resolution coronal and sagittal *T_2_*-weighted acquisitions were available. If any panel member judged the contour deviated > 1 mm from the true prostate boundary, the panel deliberated until consensus. Each slice case was reviewed at least twice. All members of the panel in attendance agreed on the final contour for each case. All four panelists were generally present for case reviews; meetings only proceeded when at least three panelists were present.

### Prostate Contouring Ground Rules

Before the consensus review, the panel decided on ground rules for situations with more than one reasonable approach. If partial volume effects were present but the prostate was clearly visible, this was included as prostate. The proximal seminal vesicles (SVs) were not intentionally included. If a visible plane divided the SVs from the prostate, the tissue was labeled as SV and excluded. Tissue with SV appearance but without a clear dividing plane on an axial slice was included as prostate. While this may lead to inclusion of a small proportion of SVs, a rule was needed, and over-estimation was preferable to undercovering the prostate. Additionally, the panel agreed this tissue would be indistinguishable from prostate on CT and was invariably included in prior trials of prostate cancer radiotherapy utilizing CT for treatment planning. The tip of the apex may not be perfectly visualized on MRI as axial slices are recommended to be 3.0 mm thick (in-plane resolution should be 0.7×0.4 mm)^10^. Therefore, at the apex, any suspicion of visible prostate, even with only subtle partial volume effects, was included as prostate.

### Auto-Segmentation Models

We invited 8 companies with commercially available prostate auto-segmentation tools to participate and 3 companies (Quibim, Radformation, and Cortechs.ai) accepted. Each commercially available tool has FDA clearance and/or CE marking for prostate auto-segmentation. Additionally, we evaluated 3 deep-learning models developed at academic centers (UCSD and Stanford). Stanford Model 1 employs a vision transformer backbone pretrained using the vision foundation model DINOv2^11^, coupled with a segmentation head. This architecture was refined through the incorporation of patch-level contrastive learning techniques (Stanford Model 2). Development and validation of the UCSD tool is described below. None of the tools were trained or previously validated using any cases in our curated reference standard dataset.

### UCSD Model Development

We collected 618 cases from 5 different datasets for training, including 92 cases from the multi-site dataset^12^, 52 cases from NCI-ISBI^13^, 139 cases from Prostate158^14^, 197 cases from Reimagine^15^, 45 cases from Cortechs.ai customer data (courtesy of Cortechs.ai), 30 cases from Prostate-3T challenge^16^, 50 cases from PROMISE12 challenge^17^, and 13 cases from The Cancer Imaging Archive (PROSTATE-DIAGNOSIS, 9 cases^18^; Fused Radiology-Pathology Prostate, 4 cases^19^). The training process utilized nnU-Net^20^, a robust and well-established model architecture. We implemented a 5-fold cross-validation strategy, with a total of 1,000 training epochs. The initial learning rate was 0.01 and gradually decreased to 0.00002 by the final epoch. Training was conducted on a single NVIDIA GeForce RTX 2080 Ti GPU, with a batch size of 2 per GPU. The input *T_2_*-weighted volumes were cropped to a patch size of 16×320×320 and resampled to a uniform voxel size of 3.0×0.4×0.4 mm³.

### Evaluation of AI tool performance

We compared each AI model segmentation to the expert consensus prostate contour of the reference standard dataset. We compared segmentation overlap using mean Dice score of the entire prostate, with 1 indicating perfect overlap and 0 indicating no overlap. We utilized a variety of clinically relevant metrics. To measure how accurately AI tools defined the prostate’s superior extent, we compared the difference in MRI slice number containing the superior-most contour between each model versus the reference standard. We repeated this to evaluate how well AI tools defined the prostate’s inferior extent. Additionally, we measured how far the AI tool strayed outside the prostate (max error outside [mm]) and cut into the prostate (max error inside [mm]). We calculated the margin required to encompass the entire prostate’s average error by comparing each auto-segmentation to a generated distance map based on the boundary of the reference standard. We determined the absolute and relative volume differences between each auto-segmentation and the reference standard, reported in mL and percentage, respectively. Since uncertainty is greatest at the superior-most and inferior-most slices, we defined the ‘main gland’ as everything but the top 2 and bottom 2 slices of the reference standard prostate. We compared each tool’s accuracy over the main gland by measuring Dice scores.

Because the purpose of this analysis is educational and not to market or disparage any commercial product, we will not label the commercial results with company names.

## Results

### Cases

We included prostate contours for 68 cases for the reference standard dataset. Cases came from 6 imaging centers, and MRI data were acquired using 3 distinct 3T scanners by two vendors (GE Healthcare; Siemens Healthineers). None of the cases used an endorectal coil. The panel reached consensus on each slice of all 68 cases. We present all slices for 2 patients to illustrate complete prostate volumes and serve as contouring guides (representative slices Figures 1-2). These are available for download as slide decks in PowerPoint.

**Figure 1.**
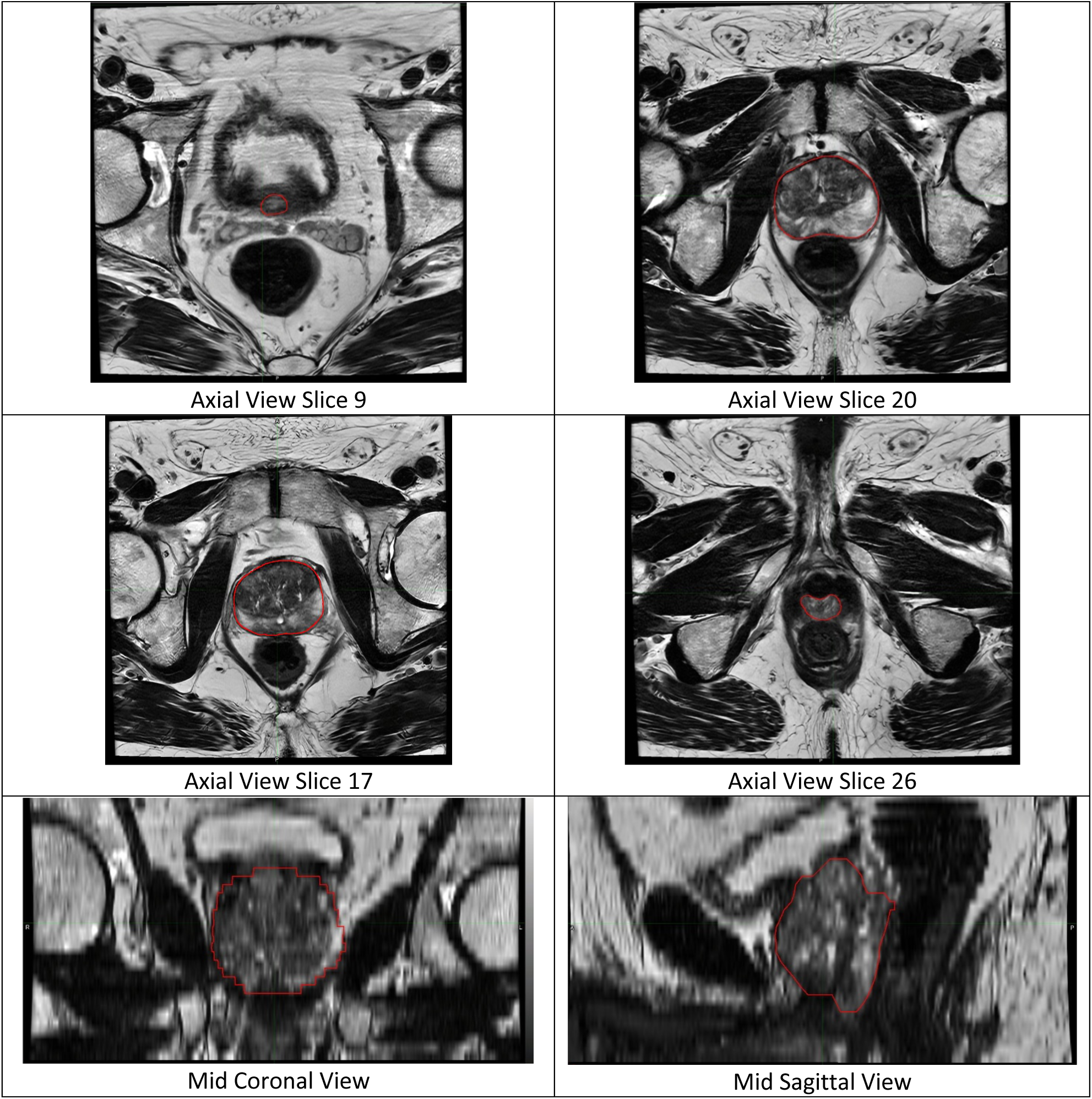
Expert-defined consensus prostate contour on MRI for a representative patient case. The expert contour is shown as red contour on axial *T_2_*-weighted slices.

**Figure 2.**
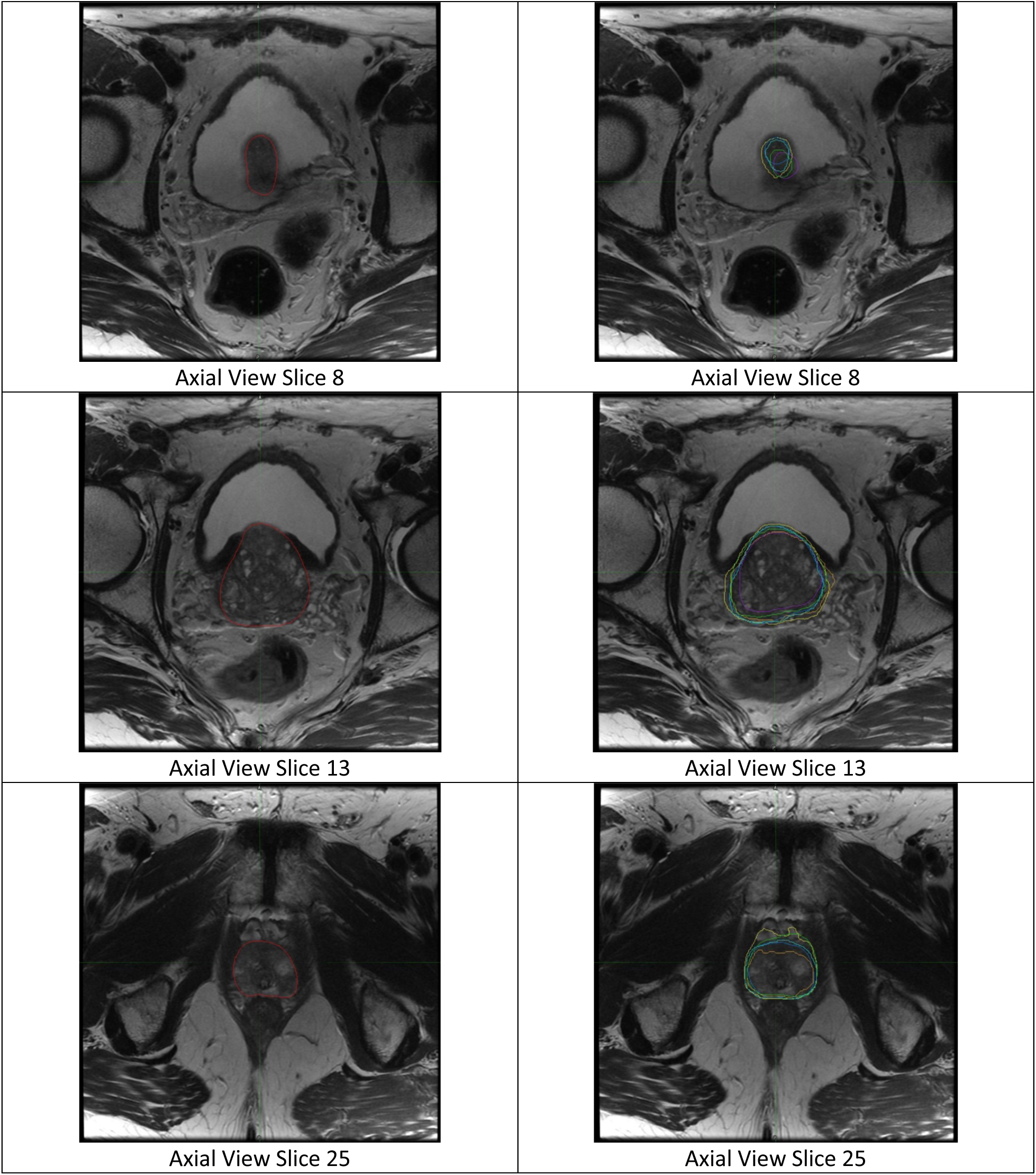
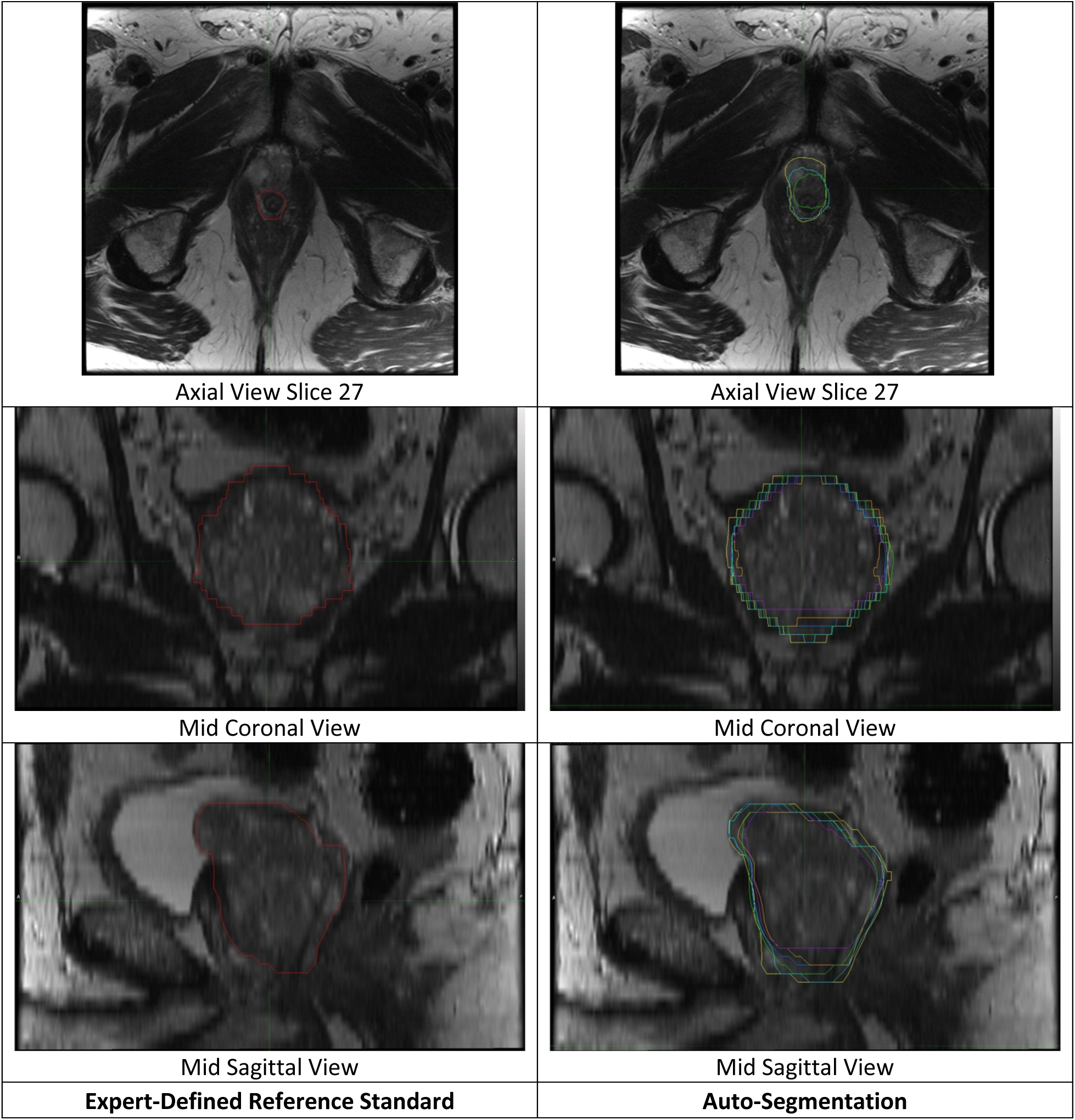
Visual comparison of expert-defined consensus contour versus all auto-segmentation models for axial *T_2_*-weighted slices from a single patient case. The left panel represents the expert-defined consensus prostate contour, shown as red contour. The right panel shows the corresponding auto-segmentation products for all models for each slice. Blue: UCSD model. Yellow: Stanford Model 1. Cyan: Stanford Model 2. Green: Company A’s product. Orange: Company B’s product. Purple: Company C’s product.

### Qualitative Observations

The greatest uncertainty regarding prostate boundary was in determining the inferior-most slice containing prostate. There was also some uncertainty at the base, though this was generally clearer than the apex. Occasionally, distinguishing the dorsal venous plexus from the prostate was difficult, especially when the plexus extended posteriorly. The panel was satisfied with the final consensus contours but recognizes there is some uncertainty, and these cases sometimes required more time to be confident the prostate boundary was accurate. Lastly, we sometimes debated what was neurovascular bundle versus prostate when determining the posterolateral prostate boundary. The most common cause of uncertainty in any part of the prostate segmentation was partial volume effects from 3.0 mm slices. On the other hand, the partial volume effects between slices amount to interpolation between slices and were generally judged to have only a small impact on the overall prostate contour shape.

### Evaluation of AI performance

All six models correctly identified the general location of the prostate in all 68 reference standard cases, except for one commercial model, which failed to generate auto-segmentations for 3 of 68 (4.4%) cases. Those 3 cases were from 2 different imaging centers. All models had greater variation at the apex and base compared to mid-gland. AI auto-segmentations are illustrated for one patient in Figure 2. For the whole prostate, median (across patients) Dice score for the six AI models ranged from 0.80 (0.72-0.85) to 0.94 (0.92-0.95) (Table 3). All six models were more accurate in the main gland with median Dice-main ranging from 0.83 (0.75-0.87) to 0.95 (0.95-0.96) (Table 3).

**Table 1.** Characteristics of the cases included in this study.

| Case Characteristics. Total Study MRI Images (n = 68) |  |  |
| --- | --- | --- |
| Cohorts |  |  |
| UC San Diego (UCSD) |  | 10 |
| Harvard University's Massachusetts General Hospital (MGH) |  | 14 |
| University of Rochester Medical Center (URMC) |  | 12 |
| UC San Francisco (UCSF) |  | 10 |
| UT Health Sciences Center San Antonio (UTHSCSA) |  | 10 |
| University of Cambridge |  | 12 |
| Cases with prominent median lobe vs. selected without regard to anatomy |  |  |
| Median Lobe |  | 23 |
| Sequential, without regard to anatomy |  | 45 |
| <u>Institution</u> | <u>MRI scanner models</u> | <u>Number of scanners</u> |
| UCSD | GE Healthcare Discovery MRI750, GE Healthcare Signa Premier | 3 |
| URMC | SIEMENS Skyra | 2 |
| MGH | GE Healthcare Signa Premier | 1 |
| UCSF | GE Healthcare Signa Premier | 2 |
| Cambridge | GE Healthcare Discovery MRI750 | 1 |
| UTHSCSA | SIEMENS Skyra | 2 |
| Total | 3 models | 11 scanners |

**Table 2.** MRI acquisition parameters *T_2_*-weighted sequences for each cohort. TR: repetition time. TE: echo time. FOV: field-of-view. FSE: fast spin echo. UCSD: University of California San Diego. MGH: Harvard University’s Massachusetts General Hospital. URMC: University of Rochester Medical Center. UTHSCSA: University of Texas Health Sciences Center San Antonio. UCSF: University of California San Francisco. Cambridge: University of Cambridge. *One of the ten patients from UCSD was scanned with a TR of 7000 ms and FOV 240 x 240 mm

| **Scanner** | **<u>UCSD\*</u>** | **<u>URMC</u>** | **<u>MGH</u>** | **<u>UCSF</u>** | **<u>Cambridge</u>** | **<u>UTHSCSA</u>** |
| --- | --- | --- | --- | --- | --- | --- |
| **Pulse sequence** | Fast Spin Echo (FSE) | Turbo Spin Echo (FSE) | Fast Spin Echo (FSE) | Fast Spin Echo (FSE) | Fast Spin Echo (FSE) | Turbo Spin Echo (FSE) |
| **TR (ms)** | 5300 | 4800 | 4800 | 2964 | 3130 | 4710 |
| **TE (ms)** | 100 | 104 | 104 | 150 | 98 | 100 |
| **FOV (mm)** | 200 x 200 | 180 x 180 | 180 x 180 | 220 x 220 | 180 x 180 | 180 x 180 |
| **Matrix [resampled dimensions]** | 320 x 320 [512 x 512] | 384 x 365 [384 x 384] | 384 x 365 [384 x 384] | 512 x 512 [320 x 320] | 448 x 256 [512 x 512] | 240 x 320 [320 x 320] |
| **Slices** | 32 | 32 | 32 | 36 | 30 | 30 |
| **Slice Thickness (mm)** | 3 | 3 | 3 | 3 | 3 | 3 |
| **Field Strength (T)** | 3 | 3 | 3 | 3 | 3 | 3 |

**Table 3.** Dice, Max error outside prostate (i.e., how far the AI segmentation strayed beyond the true prostate, in mm), Max error inside prostate (i.e., how far the AI segmentation cut into the true prostate, in mm), Average error (mm), Dice-main, Difference in superior extent of contour (number of slices), Difference in inferior extent of contour (number of slices), and Volume difference (%) of each model. The median, min, IQR, and max refer to across patients for that metric/model combination. E.g., the UCSD model gave a Dice score of 0.97 for the patient where it most accurate, 0.87 for the patient where it was least accurate, and 0.94 was the median Dice for the UCSD model across all 68 patients.

| <u>Model</u> | <u>Dice</u> |  |  | <u>Max error outside prostate (mm)</u> |  |  | <u>Max error inside prostate (mm)</u> |  |  | <u>Average error (mm)</u> |  |  |
| --- | --- | --- | --- | --- | --- | --- | --- | --- | --- | --- | --- | --- |
| Model | median |  |  | median |  |  | median |  |  | median |  |  |
|  | min | IQR | max | min | IQR | max | min | IQR | max | min | IQR | max |
| UCSD | 0.94 |  |  | 3.0 |  |  | 4.0 |  |  | 1.3 |  |  |
|  | 0.87 | 0.92-0.95 | 0.97 | 1.3 | 3.0-3.9 | 7.8 | 0.5 | 3.0-6.0 | 10.3 | 0.9 | 1.2-1.5 | 2.4 |
| Stanford Model1 | 0.89 |  |  | 7.0 |  |  | 3.0 |  |  | 2.0 |  |  |
|  | 0.75 | 0.87-0.91 | 0.94 | 3.2 | 6.0-9.0 | 15.7 | 0.6 | 2.8-4.5 | 16.7 | 1.1 | 1.7-2.2 | 4.6 |
| Stanford Model2 | 0.92 |  |  | 9.4 |  |  | 3.4 |  |  | 2.1 |  |  |
|  | 0.64 | 0.90-0.93 | 0.95 | 3.0 | 6.7-12.5 | 30.0 | 1.6 | 3.0-4.7 | 11.0 | 1.1 | 1.6-2.6 | 8.3 |
| Company A | 0.90 |  |  | 4.4 |  |  | 4.5 |  |  | 1.7 |  |  |
|  | 0.65 | 0.87-0.91 | 0.94 | 1.7 | 3.2-5.8 | 15.8 | 1.6 | 5.8-6 | 11.0 | 1.1 | 1.5-2.0 | 3.3 |
| Company B | 0.89 |  |  | 5.0 |  |  | 6.4 |  |  | 1.9 |  |  |
|  | 0.34 | 0.86-0.91 | 0.93 | 2.7 | 3.7-6.5 | 21.3 | 3.7 | 5.9-8.3 | 14.9 | 1.3 | 1.7-2.2 | 3.9 |
| Company C | 0.80 |  |  | 3.0 |  |  | 8.5 |  |  | 2.4 |  |  |
|  | 0.00 | 0.72-0.85 | 0.91 | 0.0 | 2.0-4.0 | 41.4 | 0.0 | 6.8-9.3 | 24.2 | 0.0 | 2.1-2.8 | 30.7 |
| <u>Model</u> | <u>Dice-main</u> |  |  | <u>Difference in superior extent of contour (slices)</u> |  |  | <u>Difference in inferior extent of contour (slices)</u> |  |  | <u>Volume difference (%)</u> |  |  |
| Model | median |  |  | median |  |  | median |  |  | median |  |  |
|  | min | IQR | max | min | IQR | max | min | IQR | max | min | IQR | max |
| UCSD | 0.95 |  |  | 1 |  |  | 1 |  |  | 4.7 |  |  |
|  | 0.90 | 0.95-0.96 | 0.98 | 0 | 0-1 | 2 | 0 | 0-2 | 3 | 0.1 | 3.0-8.3 | 27.1 |
| Stanford Model1 | 0.92 |  |  | 0 |  |  | 1 |  |  | 16.4 |  |  |
|  | 0.81 | 0.90-0.93 | 0.96 | 0 | 0-1 | 3 | 0 | 1-2 | 5 | 4.9 | 10.2-22.9 | 64.7 |
| Stanford Model2 | 0.94 |  |  | 1 |  |  | 3 |  |  | 4.3 |  |  |
|  | 0.72 | 0.93-0.95 | 0.96 | 0 | 0-1 | 4 | 0 | 2-4 | 9 | 0.1 | 2.3-7.7 | 36.1 |
| Company A | 0.92 |  |  | 1 |  |  | 1 |  |  | 9.2 |  |  |
|  | 0.67 | 0.89-0.93 | 0.95 | 0 | 0-1 | 3 | 0 | 0-1 | 3 | 0.1 | 4.8-17.9 | 54.5 |
| Company B | 0.91 |  |  | 1 |  |  | 1 |  |  | 12.0 |  |  |
|  | 0.36 | 0.89-0.92 | 0.95 | 0 | 1-2 | 8 | 0 | 0-1 | 5 | 0.8 | 8.1-16.8 | 62.0 |
| Company C | 0.83 |  |  | 1 |  |  | 3 |  |  | 31.4 |  |  |
|  | 0.00 | 0.75-0.87 | 0.93 | 0 | 0-2 | 4 | 0 | 2-4 | 9 | 1.6 | 22.2-43.8 | 100 |

At the prostate’s superior boundary, the auto-segmentation models differed from the reference standard by a range of 1 (0-1) to 1 (1-2) slices, median (IQR) for best and worst performing model respectively. At the inferior boundary of the prostate, the auto-segmentation models differed from the reference standard by a range of 1 (0-1) to 3 (2-4) slices, highlighting greater error in defining the inferior extent.

The best-performing model typically included at least one point 3.0 mm outside the true prostate for each patient (compared to 9.4 mm for the worst-performing model). The median (IQR across cases) maximum error outside the prostate ranged from 3.0 mm (2.0-4.0) for the best-performing model to 9.4 mm (6.7-12.5) for the worst-performing model.

The best-performing model typically excludes 3.0 mm of true prostate in at least one point somewhere in the prostate. The median (IQR) maximum error inside the prostate ranged from 3.0 mm (2.8-4.5) for the best-performing model to 8.5 mm (6.8-9.3) for the worst-performing model.

At any given point in the prostate, the expected error in either direction (inside or outside the prostate) was 1.3 mm for a typical patient for the best-performing model and 2.4 mm for a typical patient for the worst-performing model. The median (IQR) mean error ranged from 1.3 mm (1.2-1.5) to 2.4 mm (2.1-2.8).

The median (IQR) relative volume difference across patients ranged from 4.3% (2.3-7.7%) for the best-performing model to 31.4% (22.2-43.8%) for the worst-performing model (Table 3). The best-performing model typically gives estimates of prostate volume within 2.3-7.7% of the true value. The worst-performing model typically gives estimates that differ by 22-44% from the true value. Most models underestimated prostate volume (Figure 3).

**Figure 3.**
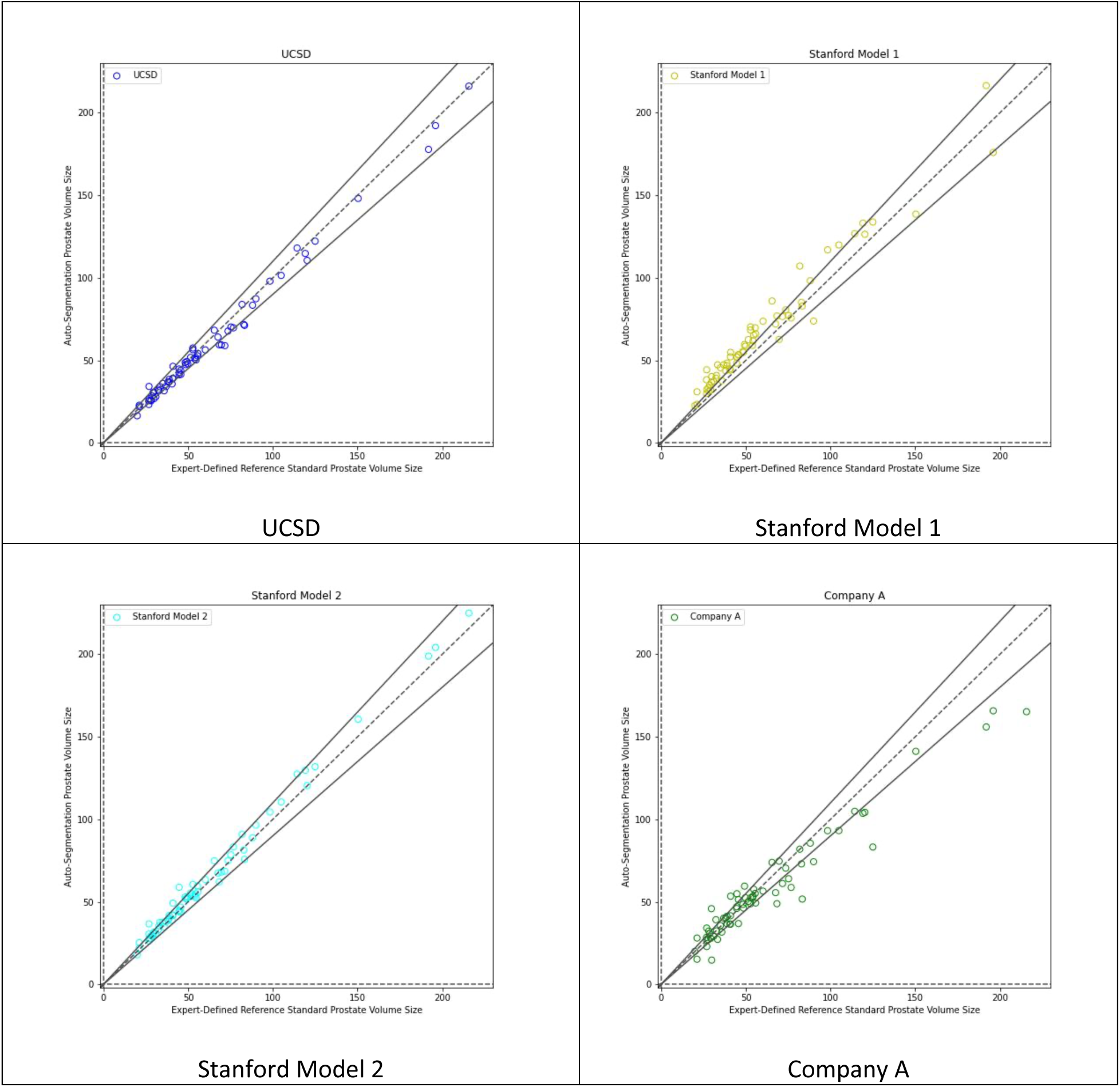
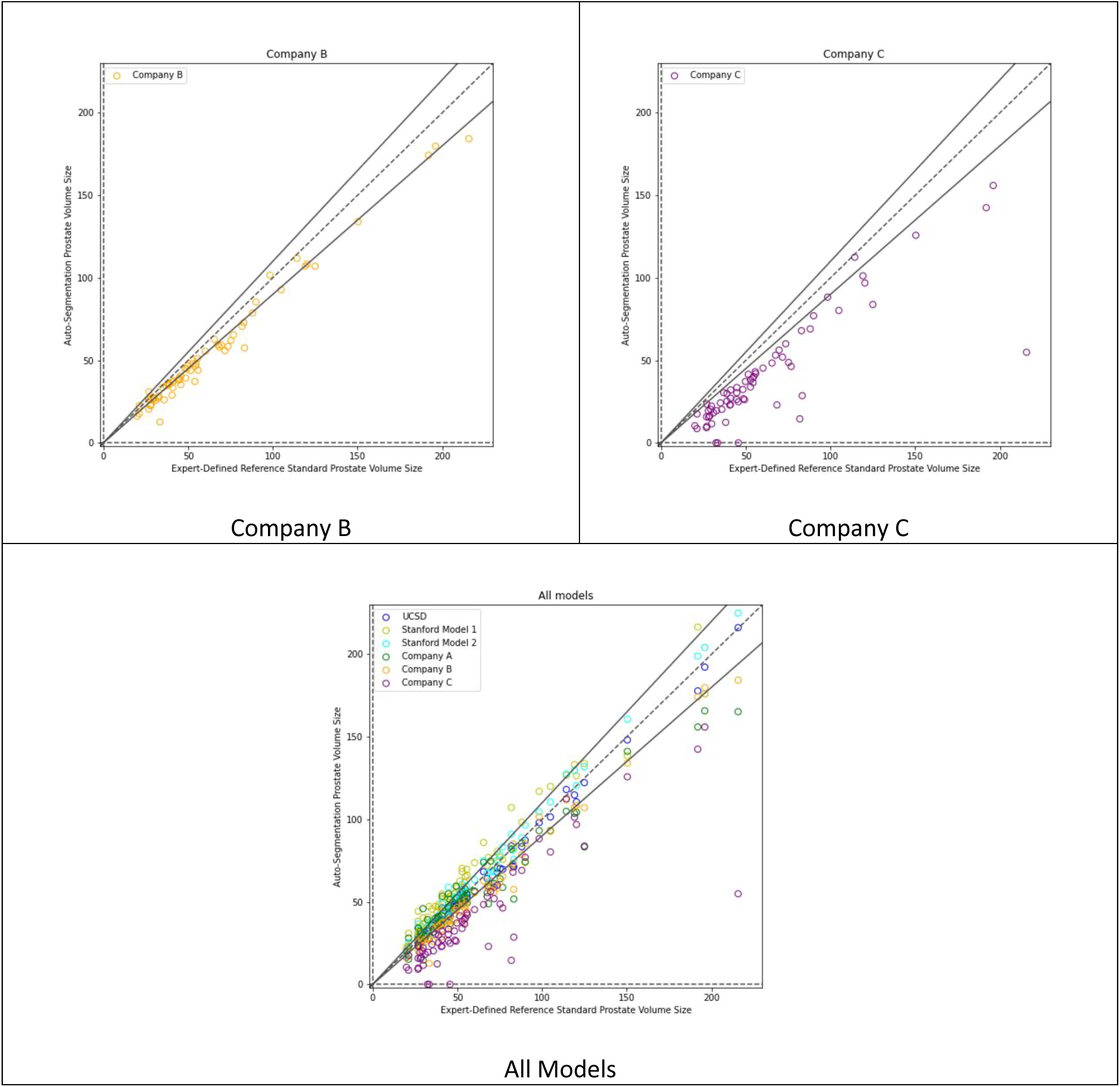
Scatter plot of auto-segmentation volume vs. expert-defined consensus contour. N=68 cases. For all panels, the X-axis shows the absolute prostate volume (mL) of the expert-defined consensus contour while the Y-axis shows the absolute prostate volume (mL) of the auto-segmentation product. If the result falls between the two solid lines, the relative volume of the auto-segmentation is within 10% (in either direction) of the expert-defined volume. Blue: UCSD model. Yellow: Stanford Model 1. Cyan: Stanford Model 2. Green: Company A’s product. Orange: Company B’s product. Purple: Company C’s product.

Presence of a prominent median lobe did not appear to be a major driver of inaccuracy for the models (Supplementary Figures 1-2 and Supplementary Tables 1-2).

Lastly, although a model’s overall performance may be quite good, there can still be considerable variation across cases. We provide representative images showing the case with lowest Dice score for each of the auto-segmentation models (Figure 4).

**Figure 4.**
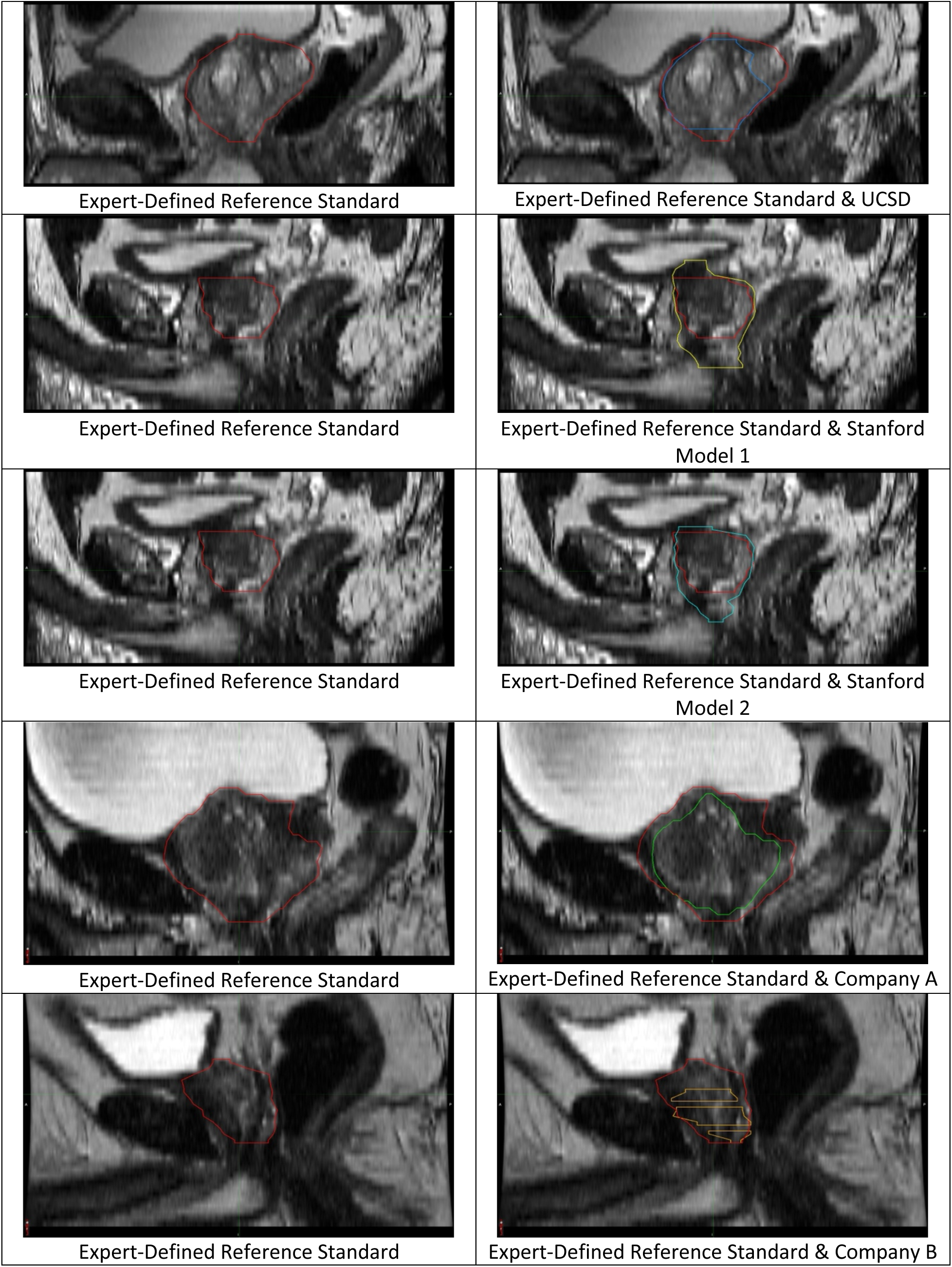
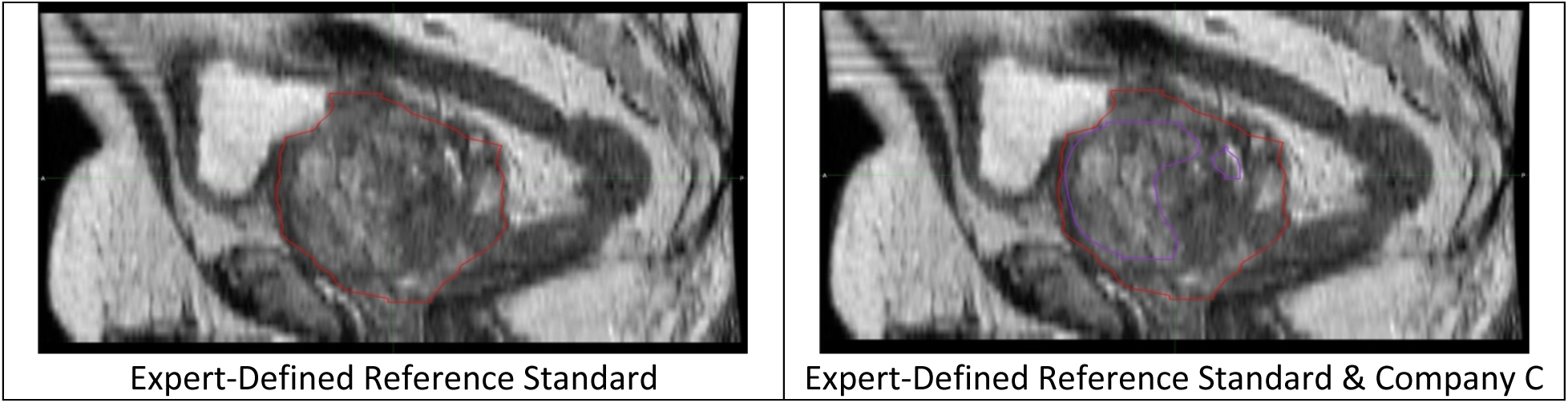
Example auto-segmentation errors. Example slices illustrating particularly bad auto-segmentation errors from the cases with the worst Dice score for each AI tool’s prostate segmentation. These are single slices from cases shown in the sagittal view. In all panels, the red contour is the expert-defined consensus contour. Blue: UCSD model. Yellow: Stanford Model 1. Cyan: Stanford Model 2. Green: Company A’s product. Orange: Company B’s product. Purple: Company C’s product.

## Discussion

Amidst a flood of published and available AI tools, the medical community needs to define the reference standard against which AI models are judged. Here, we created a reference standard dataset of 68 cases through consensus determination by an interdisciplinary expert panel for rigorous evaluation of prostate auto-segmentation AI tools. We present two cases in full as educational resources for a wide audience (radiology, radiation oncology, urology, etc.).

Using the expert consensus dataset, we tested six AI tools (three commercially available and three academic). All six models provided useful results. Prostate volume was typically adequate for calculating PSA density. However, even the best-performing models had a volume error >10% for some patients, suggesting currently these tools require physician supervision. None of the AI tools universally gave results accurate enough for radiotherapy without manual review and revision if errors >2.0 mm are considered important. The randomized MIRAGE trial compared CT-guided stereotactic body radiotherapy (SBRT) with a 4.0 mm planning margin to MRI-guided SBRT with a 2.0 mm planning margin for treatment of localized prostate cancers. Participants treated with the larger margin had significantly worse genitourinary and gastrointestinal toxicity, suggesting errors in prostate contours as small as 2.0 mm could affect outcomes^6^.

Physician contouring of the prostate shows substantial variation, and errors are common^21,22^. For radiotherapy planning, radiation oncologists have traditionally performed prostate segmentation on CT images. One study found radiation oncologists’ prostate contours were, on average, 30% larger than the true prostate volume, while still only including 84% of the prostate^23^. Another study reviewed manual contours of 300 prostates and described numerous and varied errors in each part of the prostate^22^. Here, one of the best performing AI tools had a median Dice score of 0.94, and the mean error over the full prostate contour was typically only 1.3 mm, likely clinically acceptable for radiotherapy planning and more accurate than reported accuracy of physicians contouring the prostate on CT images^24^.

Forming our reference standard was time consuming and required considerable investment by experts. We reinforce the importance of multidisciplinary collaboration in developing reference standards for meaningful comparison of AI tools. Here, the GU radiologists were experienced at reviewing high numbers of prostate MRIs and are well-trained in cross-sectional anatomy. The radiation oncologists provided the clinical context of prostate contours used for radiotherapy. Radiation oncologists strive for highly accurate contours over the full prostate in their routine clinical work, whereas approximate volumes may be acceptable for radiologists’ reporting volumes for deriving PSA density. The radiation oncologists contributed insights around seminal vesicles in the context of prostate cancer treatment, inclusion of the median lobe, and concerns of missing the tip of the apex with use of 3.0 mm MRI slices. Importantly, multidisciplinary collaboration created a better reference standard than either group would have accomplished alone. All panelists were emphatic that development of the reference standard dataset was a highly educational exercise.

According to a review of 100 commercially available AI products for medical imaging, only 36% had peer-reviewed evidence of efficacy^25^. Validating AI tools is significantly challenging but necessary to ensure patients and healthcare professionals can trust their accuracy^26^. We maintain there is great value in a reference standard dataset independent of any data used for training the AI tools it is meant to test^22,27^. This avoids inflation of accuracy due to model overfitting. Reference standard datasets should be carefully curated and only used for validation of models developed elsewhere, permitting objective head-to-head performance evaluations of existing and future AI tools. Furthermore, different AI tool outputs require different statistical evaluation methods^28^. Accuracy evaluation should include indices of overall agreement (Dice score^29^) and of clinically relevant errors (deviations from the reference standard inside and outside the prostate).

Our study has practical value for radiation oncologists and patients today. MRI-based radiotherapy planning has been shown to improve precision and may limit toxicity compared to commonly used CT-based approaches^22,30^. Delineation of the prostate on MRI is also integral to focal radiation boost for prostate cancer, which reduces cancer recurrence and metastatic spread^31,32^.

However, many radiation oncologists are unfamiliar^33^ and/or not proficient^34,35^ at contouring the prostate on MRI. Both our MRI-based consensus contouring guides (Figures 1-2) and validation of MRI-based auto-segmentation tools provide unique but related tools to facilitate widespread, accurate adoption of MRI-based radiotherapy planning.

Our study has some limitations. First, there is no purely objective way to verify the precise *in vivo* prostate boundary, so expert consensus is the best that can be achieved. Theoretically, a larger expert group could be engaged, but larger groups naturally complicate scheduling and dilute each member’s contribution. Second, acquisition protocols for prostate MRI vary across centers/scanners. We mitigated this by including data from 6 centers and 2 vendors with 3 different scanner models. We used diagnostic MRI data from PI-RADS compliant protocols rather than acquisitions designed for radiotherapy planning because this is most common. Third, although outside scope of this study, clinical use of prostate segmentation for radiotherapy has additional considerations, including (a) physiological changes to prostate position between or within treatments and (b) the need for accurate registration of MRI to CT when a separate planning CT simulation is used.

## Conclusions

We present the first expert consensus guide for prostate radiotherapy planning using MRI and a multi-institutional, interdisciplinary expert consensus dataset for meticulous evaluation of auto-segmentation AI tools. We found that some currently available AI tools are generally highly accurate, achieving average errors <2.0 mm. Physician review remains necessary, as all AI tools make clinically meaningful errors in some cases.

