## Supplemental Figure 1-2 and Table 1-2 for "Precise prostate contours: setting the bar and meticulously evaluating AI performance"

### **Supplementary Material**

A

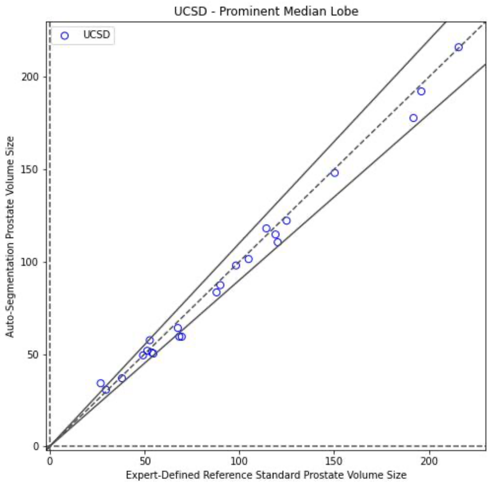


UCSD

B

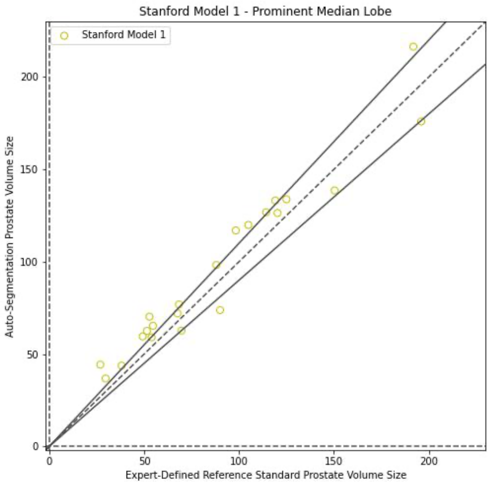


Stanford Model 1

C

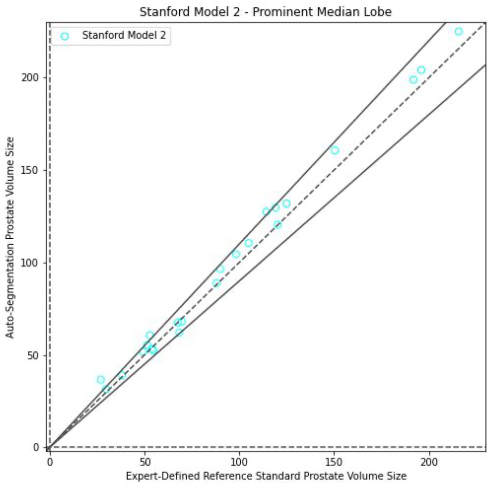


Stanford Model 2

D

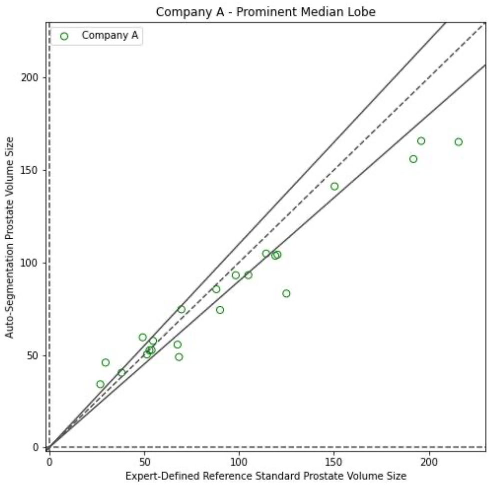


Company A

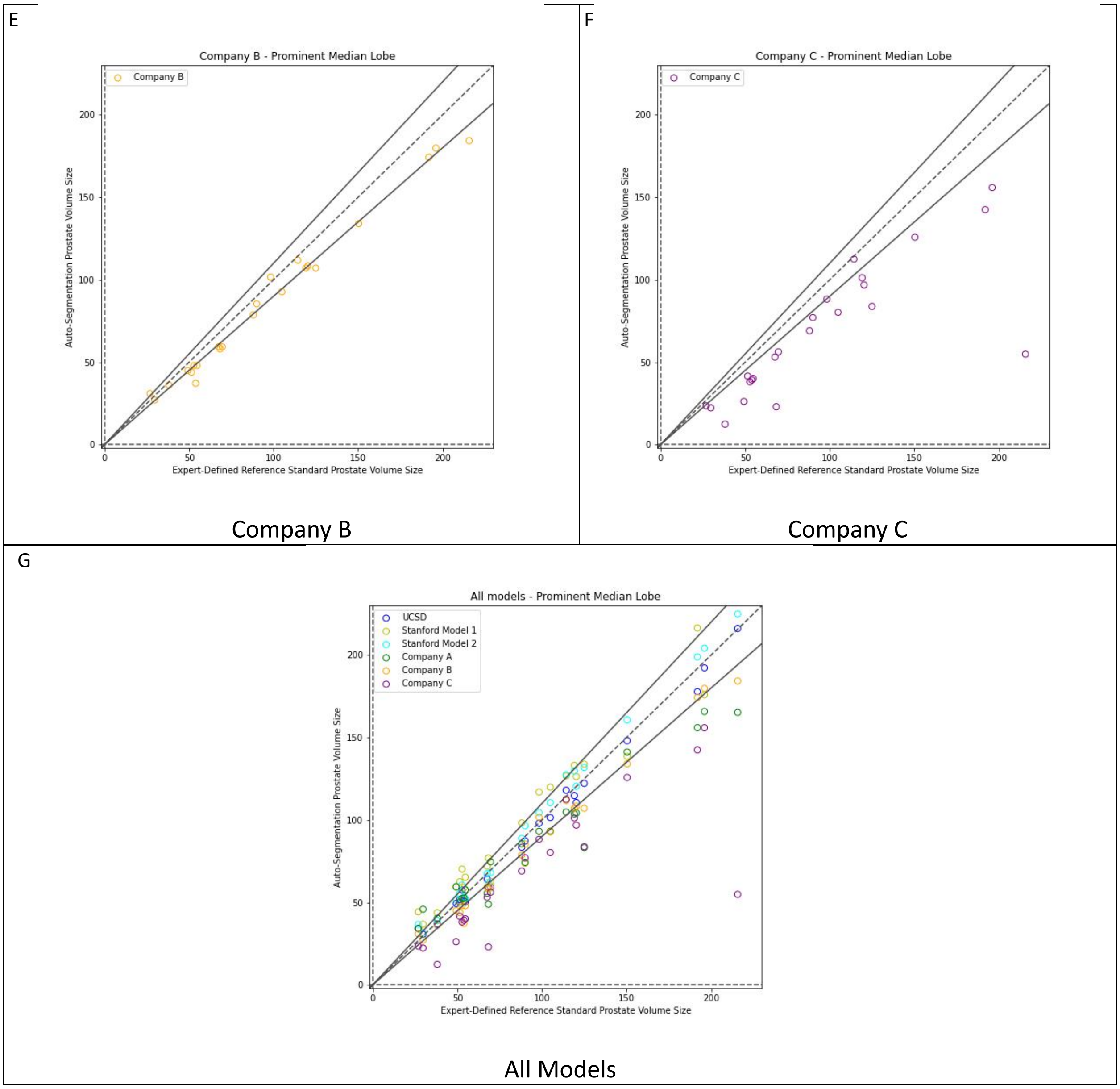


Supplementary Figure 1.

Scatter plot of auto-segmentation volume vs. expert-defined consensus contour for cases with a prominent median lobe. N=23 cases.

For all panels, the X-axis shows the absolute prostate volume (mL) of the expert-defined consensus contour while the Y-axis shows the absolute prostate volume (mL) of the auto-segmentation product for the cases specifically selected for their median lobe. Panel G compares all auto-segmentation models versus the consensus contour while panels A-F compare each model individually against the expert contour. Blue: UCSD model. Yellow: Stanford Model 1. Cyan: Stanford Model 2. Green: Company A's product. Orange: Company B's product. Purple: Company C's product.

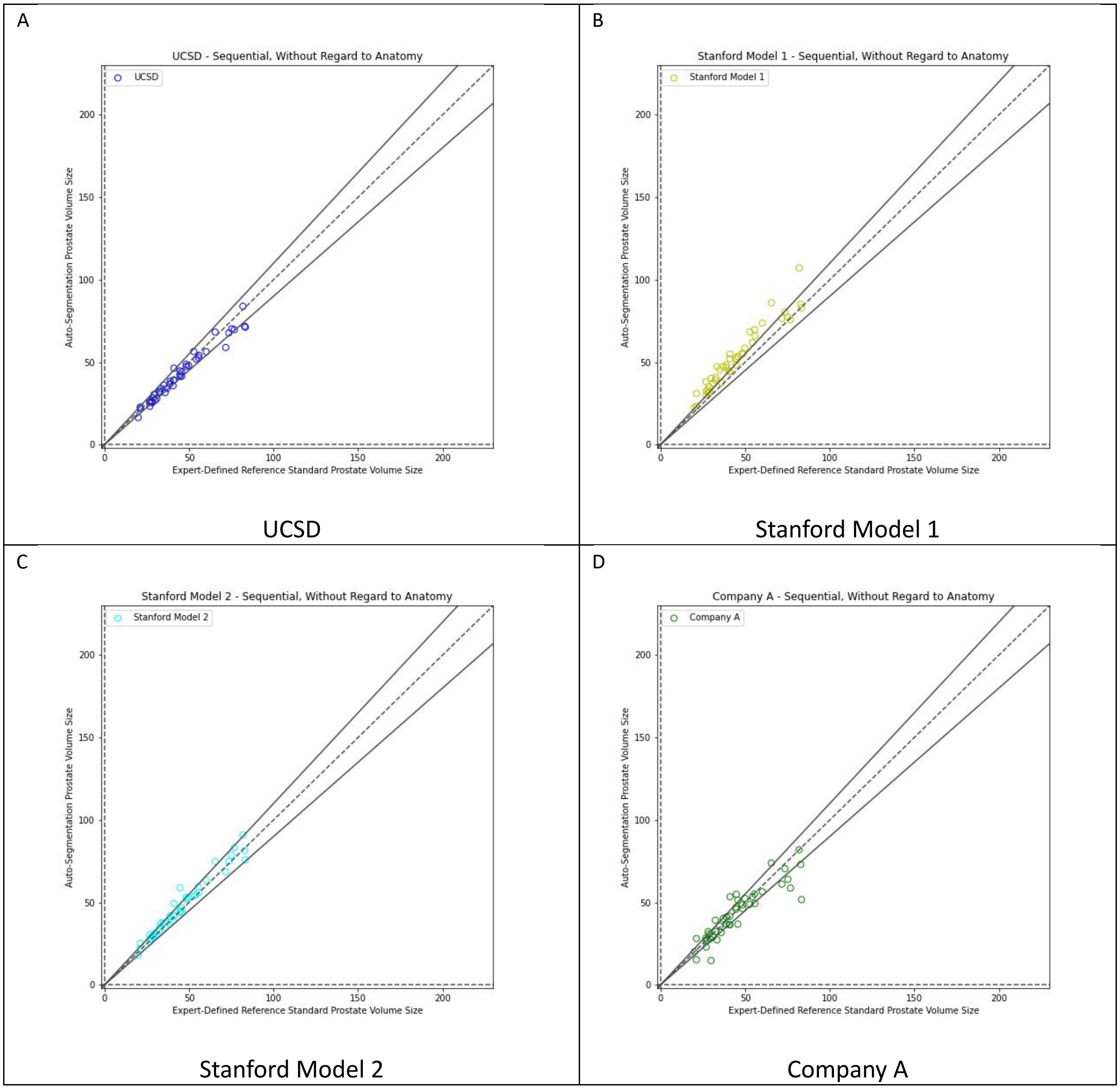
A
UCSD - Sequential, Without Regard to Anatomy
UCSD
Auto-Segmentation Prostate Volume Size
Expert-Defined Reference Standard Prostate Volume Size
0
50
100
150
200
UCSD
B
Stanford Model 1 - Sequential, Without Regard to Anatomy
Stanford Model 1
Auto-Segmentation Prostate Volume Size
Expert-Defined Reference Standard Prostate Volume Size
Stanford Model 1
C
Stanford Model 2 - Sequential, Without Regard to Anatomy
Stanford Model 2
Auto-Segmentation Prostate Volume Size
Expert-Defined Reference Standard Prostate Volume Size
Stanford Model 2
D
Company A - Sequential, Without Regard to Anatomy
Company A
Auto-Segmentation Prostate Volume Size
Expert-Defined Reference Standard Prostate Volume Size
Company A

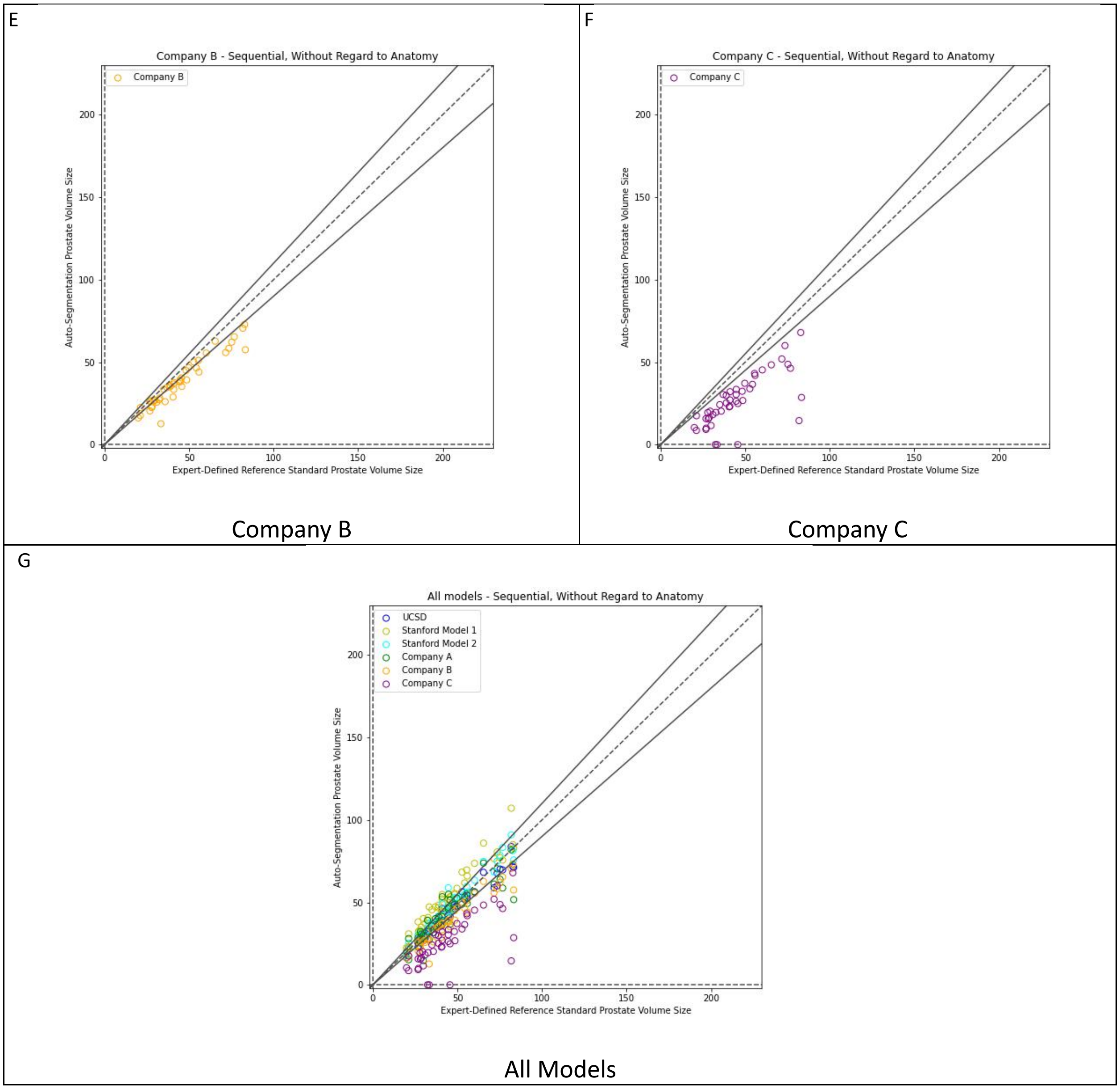


Supplementary Figure 2.

Scatter plot of auto-segmentation volume vs. expert-defined consensus contour for cases chosen without consideration of prostate anatomy. N=45 cases.

For all panels, the X-axis shows the absolute prostate volume (mL) of the expert-defined consensus contour while the Y-axis shows the absolute prostate volume (mL) of the auto-segmentation product for the cases selected without consideration of prostate anatomy. Panel G compares all auto-segmentation models versus the consensus contour while panels A-F compare each model individually against the expert contour. Blue: UCSD model. Yellow: Stanford Model 1. Cyan: Stanford Model 2. Green: Company A's product. Orange: Company B's product. Purple: Company C's product.

| Model | Dice | | | Max error outside prostate (mm) | | | Max error inside prostate (mm) | | | Average error (mm) | | |
|---|---|---|---|---|---|---|---|---|---|---|---|---|
| Model | median | | | median | | | median | | | median | | |
| | min | IQR | max | min | IQR | max | min | IQR | max | min | IQR | max |
| UCSD | 0.95 | | | 3.2 | | | 4.4 | | | 1.3 | | |
| | 0.87 | 0.94-0.96 | 0.97 | 1.9 | 3.0-5.0 | 6.8 | 0.5 | 3.5-6.0 | 10.3 | 1.0 | 1.1-1.4 | 2.4 |
| Stanford Model1 | 0.90 | | | 7.3 | | | 3.7 | | | 2.0 | | |
| | 0.75 | 0.88-0.92 | 0.94 | 4.2 | 6.0-8.5 | 15.5 | 1.5 | 3.0-9.0 | 16.7 | 1.4 | 1.8-2.3 | 4.6 |
| Stanford Model2 | 0.93 | | | 9.0 | | | 3.9 | | | 2.0 | | |
| | 0.76 | 0.92-0.94 | 0.95 | 6.0 | 6.7-15.0 | 19.4 | 1.6 | 3.0-4.8 | 11.0 | 1.4 | 1.6-2.4 | 3.7 |
| Company A | 0.90 | | | 4.4 | | | 6.0 | | | 1.7 | | |
| | 0.76 | 0.87-0.91 | 0.94 | 2.9 | 3.3-6.0 | 15.8 | 2.8 | 4.3-6.6 | 10.7 | 1.3 | 1.5-2.0 | 3.3 |
| Company B | 0.90 | | | 6.1 | | | 7.8 | | | 1.9 | | |
| | 0.79 | 0.89-0.92 | 0.93 | 3.1 | 5.0-8.5 | 12.8 | 4.0 | 6.0-9.0 | 11.2 | 1.5 | 1.7-2.2 | 3.1 |
| Company C | 0.85 | | | 3.2 | | | 0.7 | | | 2.2 | | |
| | 0.41 | 0.82-0.87 | 0.91 | 0.0 | 2.8-5.2 | 17.8 | 6.0 | 7.0-9.6 | 24.2 | 1.7 | 2.2-3.0 | 7.1 |

| Model | Dice-main | | | Difference in superior extent of Contour (slice) | | | Difference in inferior extent of contour (slice) | | | Volume difference (%) | | |
|---|---|---|---|---|---|---|---|---|---|---|---|---|
| Model | median | | | median | | | median | | | median | | |
| | min | IQR | max | min | IQR | max | min | IQR | max | min | IQR | max |
| UCSD | 0.95 | | | 0 | | | 1 | | | 3.3 | | |
| | 0.90 | 0.95-0.96 | 0.98 | 0 | 0-1 | 2 | 0 | 0-1 | 2 | 0.1 | 2.1-7.7 | 27.1 |
| Stanford Model1 | 0.92 | | | 1 | | | 1 | | | 12.5 | | |
| | 0.81 | 0.91-0.93 | 0.95 | 0 | 0-1 | 3 | 0 | 1-2 | 5 | 5.1 | 9.8-19.2 | 64.7 |
| Stanford Model2 | 0.95 | | | 1 | | | 3 | | | 5.3 | | |
| | 0.79 | 0.94-0.96 | 0.96 | 0 | 0-1 | 4 | 1 | 2-4 | 6 | 0.1 | 3.0-7.4 | 36.1 |
| Company A | 0.91 | | | 0 | | | 1 | | | 13.0 | | |
| | 0.81 | 0.89-0.93 | 0.95 | 0 | 0-1 | 3 | 0 | 0-1 | 3 | 0.6 | 5.6-19.8 | 54.5 |
| Company B | 0.92 | | | 1 | | | 1 | | | 10.6 | | |
| | 0.80 | 0.91-0.93 | 0.93 | 0 | 1-2 | 4 | 0 | 0-1 | 3 | 2.2 | 8.8-14.4 | 31.0 |
| Company C | 0.88 | | | 1 | | | 2 | | | 21.6 | | |
| | 0.41 | 0.86-0.89 | 0.93 | 0 | 1-2 | 3 | 1 | 2-3 | 5 | 1.6 | 17.8-27.5 | 75.0 |

Supplementary Table 1. Accuracy metrics for only cases with a prominent median lobe of the prostate (N=23). Difference in superior extent of contour and difference in inferior extent of contour are measured in number of slices (slices are 3 mm thick). Volume difference (%) of each model is how much the model's estimate of the prostate volume differed from the reference standard volume. The median, min, IQR, and max refer to across patients for that metric/model combination.

| Model | Dice | | | Max error outside prostate (mm) | | | Max error inside prostate (mm) | | | Average error (mm) | | |
|---|---|---|---|---|---|---|---|---|---|---|---|---|
| Model | median | | | median | | | median | | | median | | |
| | min | IQR | max | min | IQR | max | min | IQR | max | min | IQR | max |
| UCSD | 0.93 | | | 3.0 | | | 3.7 | | | 1.3 | | |
| | 0.89 | 0.92-0.95 | 0.96 | 1.3 | 3.0-3.8 | 7.8 | 1.6 | 3.0-6.0 | 7.1 | 0.9 | 1.2-1.5 | 2.1 |
| Stanford Model1 | 0.88 | | | 6.6 | | | 3.0 | | | 1.9 | | |
| | 0.81 | 0.96-0.90 | 0.94 | 3.2 | 6.0-9.0 | 15.7 | 0.6 | 2.4-3.6 | 10.4 | 1.2 | 1.7-2.2 | 3.8 |
| Stanford Model2 | 0.91 | | | 9.4 | | | 3.2 | | | 2.1 | | |
| | 0.64 | 0.90-0.93 | 0.95 | 3.0 | 6.7-12.5 | 30.0 | 1.9 | 3.0-4.7 | 8.5 | 1.1 | 1.6-2.6 | 8.3 |
| Company A | 0.89 | | | 4.3 | | | 4.3 | | | 1.6 | | |
| | 0.65 | 0.87-0.91 | 0.94 | 1.7 | 3.2-5.7 | 9.6 | 1.6 | 3.2-5.2 | 11 | 1.1 | 1.4-2.0 | 3 |
| Company B | 0.87 | | | 4.5 | | | 6.2 | | | 1.9 | | |
| | 0.34 | 0.85-0.90 | 0.93 | 2.7 | 3.6-6.0 | 21.3 | 3.7 | 5.8-8.1 | 14.9 | 1.3 | 1.8-2.2 | 3.9 |
| Company C | 0.75 | | | 3.0 | | | 8.5 | | | 2.4 | | |
| | 0.00 | 0.71-0.82 | 0.87 | 0.0 | 1.5-3.6 | 41.4 | 0.0 | 6.8-9.3 | 14.7 | 0.0 | 2.1-2.8 | 30.7 |
| **Model** | **Dice-main** | | | **Difference in superior extent of contour (slices)** | | | **Difference in inferior extent of contour (slices)** | | | **Volume difference (%)** | | |
| Model | median | | | median | | | median | | | median | | |
| | min | IQR | max | min | IQR | max | min | IQR | max | min | IQR | max |
| UCSD | 0.95 | | | 1 | | | 1 | | | 4.8 | | |
| | 0.92 | 0.94-0.96 | 0.97 | 0 | 0-1 | 2 | 0 | 1-2 | 3 | 0.1 | 3.3-9.3 | 17.8 |
| Stanford Model1 | 0.92 | | | 0 | | | 1 | | | 17.7 | | |
| | 0.86 | 0.90-0.93 | 0.96 | 0 | 0-1 | 2 | 0 | 1-2 | 4 | 4.9 | 10.6-25.5 | 45.9 |
| Stanford Model2 | 0.94 | | | 1 | | | 3 | | | 4.2 | | |
| | 0.72 | 0.93-0.95 | 0.96 | 0 | 0-1 | 4 | 0 | 2-4 | 9 | 0.5 | 2.0-8.1 | 31.8 |
| Company A | 0.92 | | | 1 | | | 1 | | | 7.4 | | |
| | 0.67 | 0.90-0.93 | 0.95 | 0 | 0-1 | 2 | 0 | 0-1 | 3 | 0.1 | 4.2-14.9 | 50.7 |
| Company B | 0.90 | | | 1 | | | 1 | | | 13.8 | | |
| | 0.36 | 0.88-0.92 | 0.95 | 0 | 45293 | 8 | 0 | 0-1 | 5 | 0.8 | 7.7-18.7 | 62.0 |
| Company C | 0.78 | | | 1 | | | 3 | | | 35.8 | | |
| | 0.00 | 0.74-0.86 | 0.89 | 0 | 0-2 | 4 | 0 | 2-4 | 9 | 17.5 | 26.1-45.0 | 100 |

Supplementary Table 2. Accuracy metrics for only cases selected at random, without regard to anatomy (N=45). Difference in superior extent of contour and difference in inferior extent of contour are measured in number of slices (slices are 3 mm thick). Volume difference (%) of each model is how much the model's estimate of the prostate volume differed from the reference standard volume. The median, min, IQR, and max refer to across patients for that metric/model combination.
